# Smart Glasses for Gait Analysis in Parkinson’s Disease: A preliminary study

**DOI:** 10.1101/2022.10.22.22281214

**Authors:** Ivana Kiprijanovska, Simon Stankoski, Martin Gjoreski, James Archer William Archer, John Broulidakis, Ifigeneia Mavridou, Bradley Hayes, Charles Nduka, Hristijan Gjoreski

## Abstract

Parkinson’s disease (PD) is one of the most common neurodegenerative disorders of the central nervous system, which predominantly affects patients’ motor functions, movement, and stability. Monitoring movement in patients with PD is crucial for inferring motor state fluctuations throughout daily life activities, which aids in disease progression analysis and assessing how patients respond to medications over time. In recent years, there has been an increase in the usage of wearable sensors for PD symptom monitoring. In this study, we present a preliminary analysis of smart glasses equipped with IMU sensors to provide objective information on the motor state in patients with PD. Data were collected from seven Parkinson’s patients with varying levels of symptom severity. The patients performed the Timed-Up-and-Go (TUG) Test while wearing IMU-equipped glasses. Our analysis indicates that smart glasses can provide information about patients’ gait that can be used to assess the severity level of the PD as measured by two standardized questionnaires. Furthermore, patient-specific clusters can be easily detected in the sensor data, hinting at the development of personalized models for patient-specific monitoring of symptom progression. Therefore, smart glasses have the potential to be used as an unobtrusive and continuous screening tool for PD patients’ gait, enhancing medical assessment and treatment.

CCS CONCEPTS • Applied computing • Life and medical sciences • Health informatics

**ACM Reference Format:** First Author’s Name, Initials, and Last Name, Second Author’s Name, Initials, and Last Name, and Third Author’s Name, Initials, and Last Name. 2022. The Title of the Paper: ACM Conference Proceedings Manuscript Submission Template: This is the subtitle of the paper, this document both explains and embodies the submission format for authors using Word. In Woodstock ‘18: ACM Symposium on Neural Gaze Detection, June 03–05, 2018, Woodstock, NY. ACM, New York, NY, USA, 10 pages. NOTE: This block will be automatically generated when manuscripts are processed after acceptance.

## 1 INTRODUCTION

Parkinson’s disease (PD) is one of the most common progressive neurological disorders. In the past 25 years, the prevalence of Parkinson’s disease has doubled globally. Global estimates in 2019 showed over 8.5 million individuals living with PD. PD predominantly impairs patients’ motor abilities but is also associated with a wide variety of non-motor complications, including cognitive impairment, mental health and sleep disorders, sensory disturbances, and other behavioral problems. The severity and frequency of the symptoms usually increase as the disease progresses over time, impacting the patient’s mental health and self-esteem, and significantly worsening their quality of life. The progression of the motor dysfunctions, as the most common and disabling symptoms of PD, requires monitoring of patients’ gait over time. Monitoring and analysis of PD patients’ gait are critical for early disease diagnosis and quantifying disease progression. For these reasons, having efficient and reliable tools for gait analysis is essential.

The Timed-Up-and-Go (TUG) test is a clinical tool commonly used for measuring motor dysfunction in PD. The subject will stand up from sitting, walk 3 meters, turn, walk back to the chair and sit back down, and the examiner will measure, in seconds, the time taken to complete the task [1]. Total time taken has been shown to correlate with disease severity [2], risk of falls over the next year [3][4], and response to dopamine-based therapies [5]. The TUG test is limited in that it measures the patient at a single point and is not truly representative of all activities of daily living (ADLs). In addition, dopaminergic medications have “on” phases (peak effect) and “off” phases (trough effect), which makes the comparison of TUG testing over time challenging as patients may be in a different part of this cycle [6].

Due to this, many studies have looked at wearable devices which can track movements in PD without the need for a direct observer. Multiple studies have validated wearable devices during TUG testing and found reliability in measuring the total time taken [7] and predicting fall risk and gait and mobility parameters [8]. Wearable sensors have also been shown to delineate the time taken for specific movements such as the sit-to stand or stand-to-sit transition [9], gait speed, and turning speed [10]. A study by Weiss et al. [11] measured data from a sensor worn on the lower back for 3 days and found a significantly improved ability to predict fall risk compared to commonly used clinical scoring measures. Wearable sensors have also been used to measure freezing of gait (FOG) symptoms, where the patient is unable to initiate movement, over longer time periods [12][13]. This is important as there may be specific triggers for this, such as walking through a narrow space, which cannot be replicated during a TUG test.

Therefore, wearable devices can be used to enhance PD disease monitoring and aid medication decisions by giving clinicians information about the patient in a wider variety of settings and over a longer time period. In this paper, we present a preliminary analysis of the ability of smart glasses equipped with Inertial Measuring Unit (IMU) sensors to provide objective information on the motor state in patients with PD.

## 2 DATA

For the analysis, a total of 7 PD patients (4 females and 3 males, with a mean age of 77 ± 7.3, range 68–87) with different levels of disease symptom severity were recruited. As per ethical requirements, all participants provided written informed consent before participating in the study. For data collection, we used emteq’s OCOSense™ smart glasses (Figure 1). The smart glasses are equipped with: (i) seven OCO™ sensors that measure skin movement in three directions (located at the green rectangles in Figure 1), and (ii) a 9-axis inertial sensor (accelerometer, gyroscope, and magnetometer), including an altimeter (located in the right arm of the glasses frame, indicated by the purple rectangle in Figure 1). The orientation of the inertial sensor in the glasses can be determined by aligning the black dot on the zoomed inertial sensor in Figure 1 and the white dot below the purple rectangle on the glasses frame. This results in the x axis being vertically oriented (opposite of the gravitational force), the y axis being aligned with the glasses arm (pointing towards the ear), and the z axis being the horizontal axis (pointing to the head).

**Figure 1.**
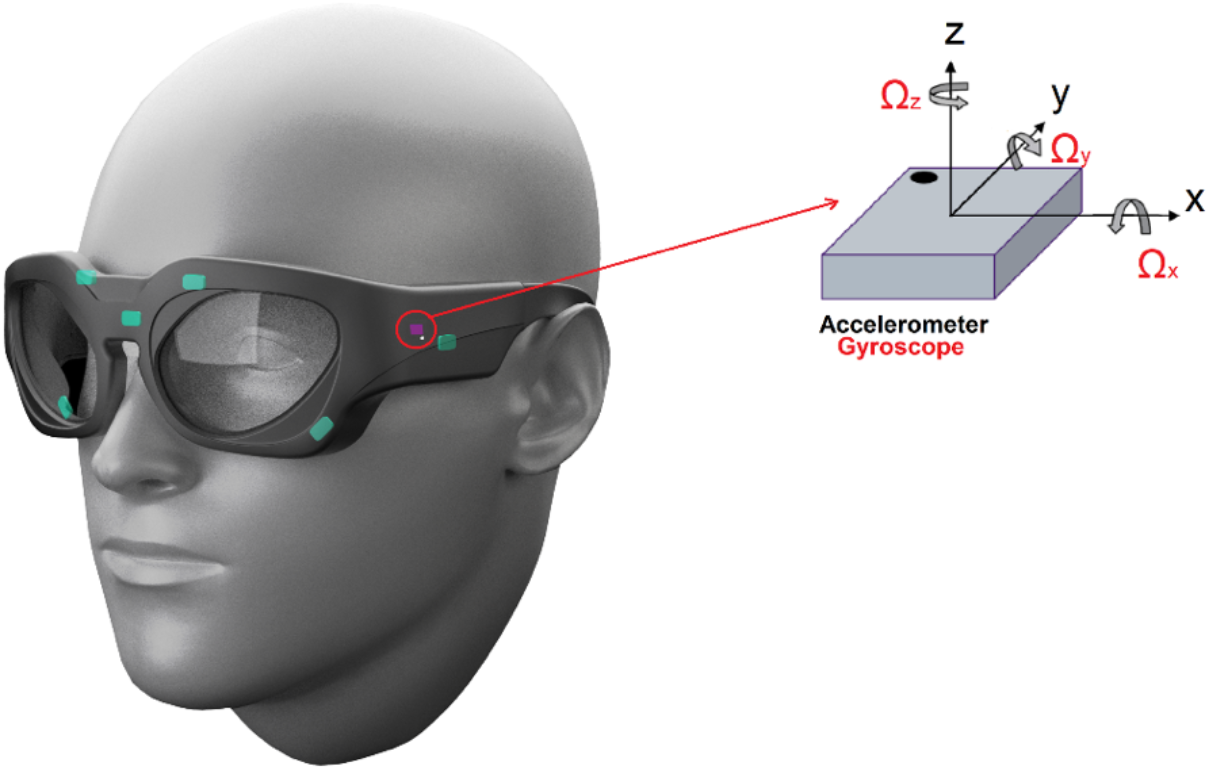
emteq’s OCOSense™ glasses and the sensors’ location. The green rectangles represent the OCO™ sensors, and the purple rectangle represents the 9-axis inertial sensor.

The experimental protocol was based on the Timed-Up-and-Go (TUG) Test: the participants sat on a chair, stood up, walked straight for 3 meters at their normal speed, made a 180° turn around an obstacle, walked straight back to the chair, did another 180° turn, and finally sat down on the chair. They performed a total of 5 sessions of the TUG Test. All participants were able to perform all sessions independently, without an assistive device. The sessions were also recorded with a video camera. The study was reviewed and approved by the NHS research ethics committee, (ref: 18/WM/0205) and took place at Queen Victoria Hospital in East Grinstead, England.

All participants also completed two questionnaires: the Freezing of Gait (FOG) Questionnaire and the Parkinson’s Disease Questionnaire (PDQ-8). The former includes 6 questions and is focused on FOG severity and gait impairments over the last week, while the latter includes 8 questions about mobility, activities of daily living, emotional well-being, stigma, social support, cognition, communication, and bodily discomfort, and is used to quantify the quality of life among PD patients. Each question was scored between 0 and 4. A higher score on the FOG questionnaire corresponds to more severe FOG episodes. A higher score for the PDQ-8 questionnaire signifies a poorer quality of life and a more severe form of the disease. The final scores from the questionnaires, alongside the participant demographics, are presented in Table 1. The questionnaire’s score for each participant was divided by the total possible score (for each questionnaire separately), and the final scores in the table are given as a percentage out of 100.

**Table 1:** Demographics data and questionnaires’ scores.

| Participant | Age range | Sex | FOG Score (%) | PDQ-8 Score (%) |
| --- | --- | --- | --- | --- |
| 1 | 66-70 | Male | 20.8 | 46.9 |
| 2 | 71-75 | Female | 4.2 | 40.6 |
| 3 | 81-85 | Female | 87.5 | 75.0 |
| 4 | 66-70 | Female | 66.7 | 37.5 |
| 5 | 71-75 | Male | 25.0 | 31.3 |
| 6 | 85-89 | Male | 66.7 | 65.6 |
| 7 | 85-89 | Female | 33.3 | 34.4 |

The TUG Test is considered a reliable and valid test for assessing general functional mobility [1][2]. Patients with PD experience changes in their gait, including a decrease in walking speed and reduced step length and pattern. This is demonstrated by the duration and the number of steps required by the patients to complete the TUG Test. These values were obtained by analyzing the video recordings and are presented in Table 2 and Table 3.

**Table 2:** Duration of walking (in seconds) by each participant during the 5 Timed-Up-and-Go (TUG) sessions.

| Participant | Session 1 | Session 2 | Session 3 | Session 4 | Session 5 | Mean ± Std | FOG Score | PDQ-8 Score |
| --- | --- | --- | --- | --- | --- | --- | --- | --- |
| 1 | 19 | 19 | 18 | 18 | 20 | $18.8 \pm 0.7$ | 20.8 | 46.9 |
| 2 | 12 | 11 | 12 | 12 | 14 | $12.2 \pm 1.0$ | 4.2 | 40.6 |
| 3 | 30 | 23 | 23 | 22 | 21 | $23.8 \pm 3.2$ | 87.5 | 75.0 |
| 4 | 12 | 13 | 13 | 13 | 13 | $12.8 \pm 0.4$ | 66.7 | 37.5 |
| 5 | 10 | 10 | 10 | 10 | 10 | $10.0 \pm 0.0$ | 25.0 | 31.3 |
| 6 | 21 | 17 | 17 | 17 | 19 | $18.2 \pm 1.6$ | 66.7 | 65.6 |
| 7 | 14 | 13 | 13 | 12 | 12 | $12.8 \pm 0.7$ | 33.3 | 34.4 |

**Table 3:** Number of steps performed by each participant in the 5 Timed-Up-and-Go (TUG) sessions.

| Participant | Session 1 | Session 2 | Session 3 | Session 4 | Session 5 | Mean ± Std | FOG Score | PDQ-8 Score |
| --- | --- | --- | --- | --- | --- | --- | --- | --- |
| 1 | 16 | 15 | 16 | 14 | 15 | $15.2 \pm 0.7$ | 20.8 | 46.9 |
| 2 | 15 | 14 | 14 | 14 | 14 | $14.2 \pm 0.4$ | 4.2 | 40.6 |
| 3 | 32 | 32 | 34 | 32 | 34 | $32.8 \pm 1.0$ | 87.5 | 75.0 |
| 4 | 15 | 16 | 16 | 16 | 16 | $15.8 \pm 0.4$ | 66.7 | 37.5 |
| 5 | 11 | 11 | 11 | 11 | 11 | $11.0 \pm 0.0$ | 25.0 | 31.3 |
| 6 | 19 | 20 | 20 | 20 | 20 | $19.8 \pm 0.4$ | 66.7 | 65.6 |
| 7 | 16 | 16 | 15 | 16 | 16 | $15.8 \pm 0.4$ | 33.3 | 34.4 |

The results show that participant 3, who has the most severe PD symptoms according to the scores of the questionnaires (FOG Score = 87.5, PDQ Score = 75.0), shows notably increased time for completion of the TUG Test (23.8 ± 3.2 seconds) and an increased number of steps (32.8 ± 1.0), compared to the rest of the participants. Participant 5, who needed the least time on average to complete the 5 sessions of the TUG Test (10.0 ± 0.0 seconds) and made the smallest number of steps (11.0 ± 0.0) is ranked 5th on the FOG Score scale and 7th on the PDQ-8 scale, indicating that this participant is the one with the least severe form of the disease.

To further determine the strength of the relationship between the questionnaires’ scores and the duration and number of steps performed during the TUG Test, we performed linear regression analysis and obtained the correlation coefficients. These results are depicted in Figure 2Figure 2.

**Figure 2:**
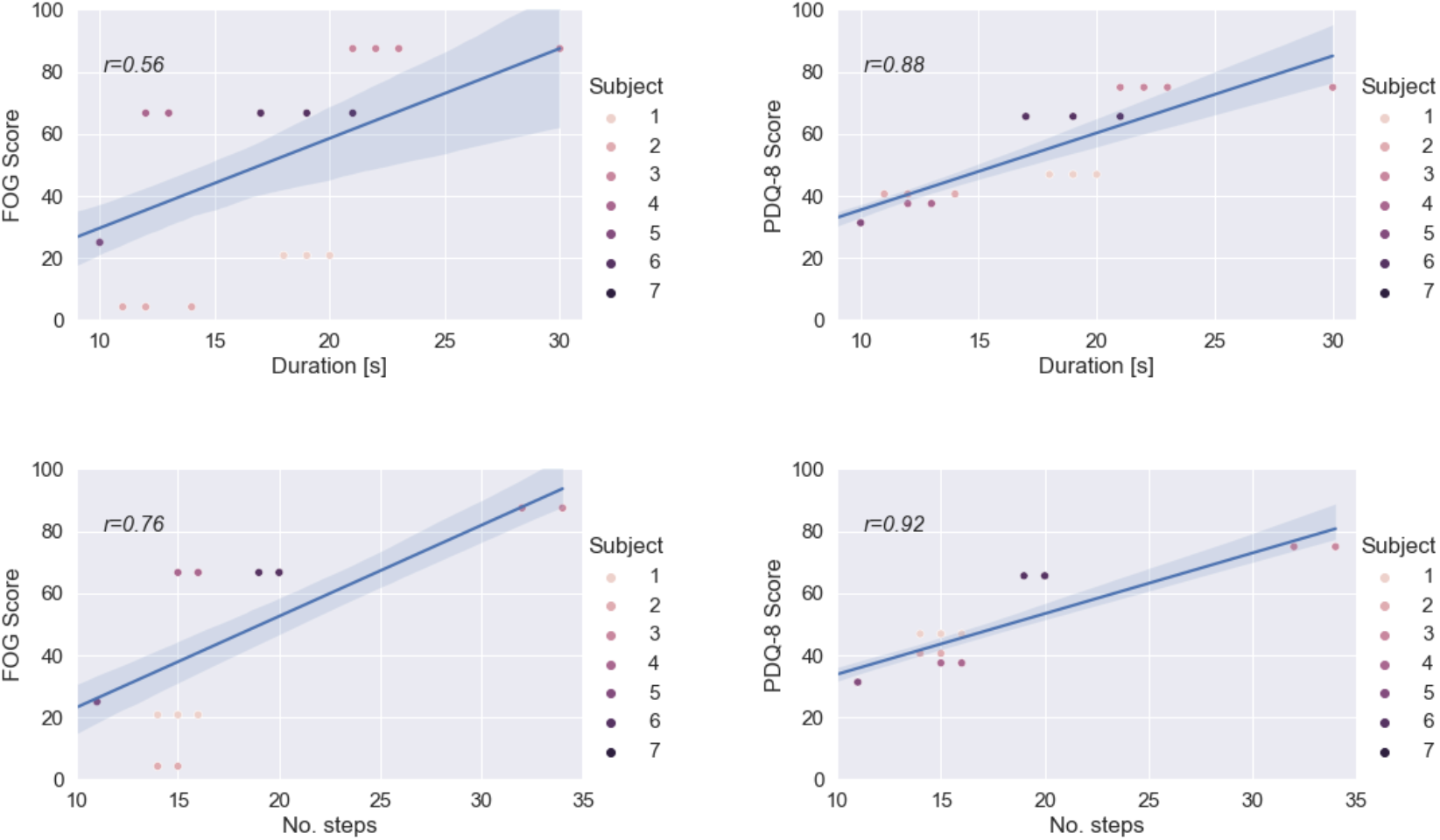
Linear Regression Analysis of the FOG (first column figures) and PDQ-8 (second-columns figures) Questionnaires versus duration (first-row figures) and number of steps (second-row figures) performed during the TUG Test.

A positive correlation was observed in all four tested cases. In general, the results show that the duration and the number of steps required for the TUG Test are stronger correlated with the PDQ-8 Score that the FOG Score. The strongest correlation was observed between the number of steps performed during the TUG Test and the PDQ- 8 Score (r=0.92).

Moreover, we also analyzed the strength of the linear relationship between the age of the patients and their questionnaires scores (FOG and PDQ-8 score). In this case, we also observed a positive correlation for both scores, with PDQ-8 score showing a greater correlation with the age of the patients (r=0.48) than the FOG score (r=0.38).

## 3 SENSOR DATA PROCESSING AND ANALYSIS

The sensor data were collected using emteq’s OCOSense™ smart-glasses that provided 3-axis accelerometer, 3-axis gyroscope, and 3-axis magnetometer data, all sampled at 50 Hz. For the sensor data analysis, we only used the accelerometer and gyroscope data. Additionally, we calculated the magnitude of both sensors, which resulted in eight sensor streams for the analysis, overall.

A 3rd order Butterworth lowpass filter was applied to extract the required frequency components from the IMU signals, reduce the high-frequency artifacts, and preserve the gait information within the signals. After the filtering steps, the data were segmented using a sliding window of 2 seconds and a 50% overlap. Finally, we extracted 9 general-purpose statistical features from each of the sensor signals, resulting in a total of 72 features. The features included the mean, standard deviation, minimum, maximum, kurtosis, skewness, value range, root mean square, and interquartile range. We conducted the initial analysis in this study with a small number of simple features that are computationally inexpensive in an effort to provide real-time processing on the glasses in the future.

To understand how the motion data coming from the sensors (recorded during the TUG Test) is related to the tests scores obtained from the questionnaires, we performed analysis using the SHAPley Additive exPlanations (SHAP) method. More precisely, we fitted a machine learning model based on the Extreme Gradient Boosting algorithm (XGBoost) to the obtained feature set, where the target variables were either the FOG or the PDQ-8 Score. The SHAP method computes the contribution of each feature to the model’s output, so we were able to identify if any of the features have a meaningful impact on the model’s output. The results of the SHAP-based analysis are shown in Figure 3.

**Figure 3:**
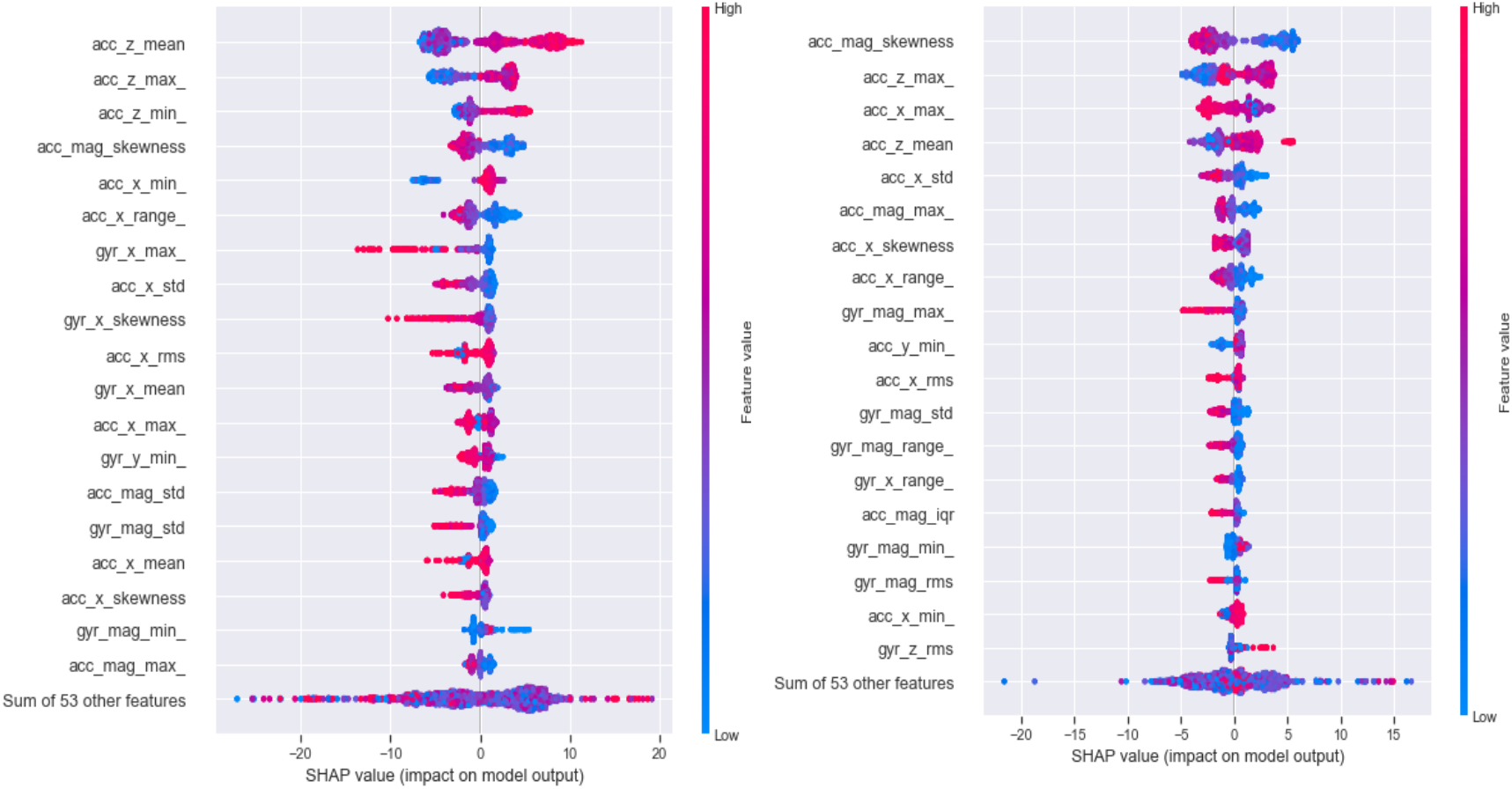
SHAP feature importance, measured as the mean absolute Shapley values. FOG Score (left), PDQ-8 Score (right).

The figure shows the top 19 most informative features sorted in descending order, for both the FOG Score (on the left) and the PDQ-8 Score (on the right). The SHAP values show in which direction the features are contributing to the target variable, i.e., the x-axis shows whether the effect of the feature value is associated with higher or lower model output, or in our case, a higher or lower FOG/PDQ-8 score. Additionally, the color (from blue to red) provides information about the original value of the features. Blue represents lower feature values and red represents higher feature values. This allows us to identify whether the features have a positive or negative correlation with the model’s output.

The figure on the left (Figure 3, FOG Score) indicates that the top three features are the mean, the maximum and the minimum values of the accelerometer’s z-axis. Higher values on these three features correspond to a higher model output (FOG Score), which suggests that greater acceleration along the z axis is associated with more severe FOG symptoms. In the context of the sensor position, the conclusion that can be derived from this is that the features associated with lateral head movements (the z axis of the accelerometer in the glasses is the horizontal axis pointing to the head, see Figure 1) have the greatest influence on the output of the model. In other words, greater acceleration of the head in lateral directions may indicate a higher degree in the disease severity (as measured by the FOG questionnaire). We speculate that this might be the result of patients’ less controlled steps and movements as their PD symptoms worsen. Those could cause unsteadiness of the body and occurrence of involuntary movement that cause higher acceleration even in the lateral head direction. However, given the relatively small data sample used for this analysis, there is also a possibility that the head and overall body posture of the patients included in the study contribute to these results. The fourth feature is the skewness of the acceleration’s magnitude, and it can be observed that higher skewness is related to lower FOG scores. Given that the skewness is a measure of symmetry in the distribution of the values, this feature can be interpreted as the less-symmetrical distribution of the acceleration’s magnitude is related to more severe PD symptoms (as measured by the FOG scores). This may be because more severe symptoms cause less controlled movements, which cause distribution shifts. From the rest of the features, it is apparent that the features extracted from the x-axis are inversely related to the model’s output (FOG Score), i.e., higher feature values correspond to a lower FOG Score.

From the figure on the right (Figure 3, PDQ-8 Score), it can be determined that the top-ranked feature (acceleration magnitude) is the same as the fourth-ranked feature from the left figure (FOG Score). The same relation can also be noted, i.e., higher skewness is related to a lower PDQ-8 Score. Furthermore, the second and the fourth-ranked features are related to the accelerometer’s z-axis, and similarly as with the FOG Score, higher values in these features correspond to a higher model output (PDQ-8 Score).

Lastly, we used the t-distributed stochastic neighbour embedding (t-SNE) method [14] to visualize the data and to determine whether the sensor data recorded from the glasses could reveal additional information about the patients. For this analysis, we only used the feature vectors extracted from the walking segments from the TUG Test sessions. The result is shown in Figure 4. The figure shows seven clusters of data, which indicates that all patients have a unique or separable walking style. The walking style may also depend on the severity of the PD. These results may also indicate that the glasses-based data is sensitive enough to develop personalized models for patient-specific monitoring of symptom progression.

**Figure 4:**
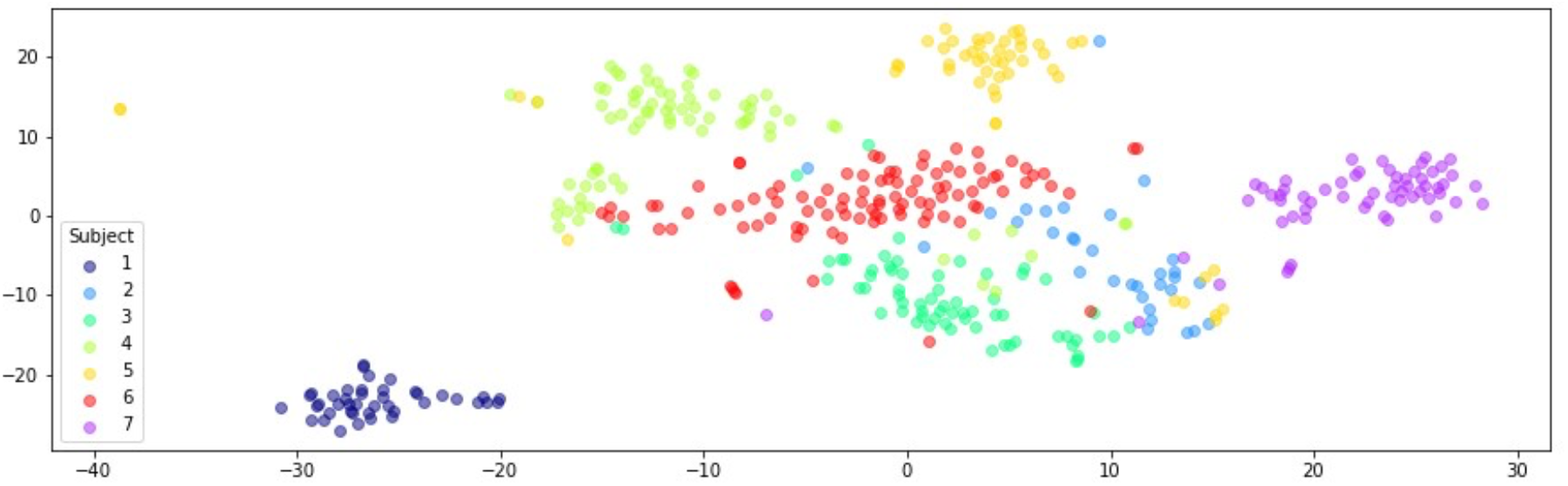
t-SNE visualization of the walking data from the Timed-Up-and-Go Test.

## 4 CONCLUSION

In this paper, a preliminary analysis was presented with regards to the ability of emteq’s novel OCOSense™ smart glasses equipped with IMU sensors to provide objective information on the motor state in patients with Parkinson’s disease. We used data from seven Parkinson’s patients with varying levels of the disease’s symptoms severity who were performing the Timed-Up-and-Go (TUG) Test while wearing the glasses. Although only a small group of PD patients was examined in the experiment, the preliminary analysis suggests that IMU-equipped smart glasses have the potential to provide information about patients’ gait and can be used to assess the severity level of Parkinson’s disease as measured by two standardized questionnaires. They can therefore be considered as a screening tool that will continuously monitor Parkinson’s disease patients’ gait and motor activity. Moreover, the analysis of the walking data has shown that the patients have a unique or separable walking style, which may also depend on the severity of the disease. This indicates that the glasses-based sensor data is sensitive enough to develop personalized models for patient-specific monitoring of symptom progression.

Future work will include the acquisition and analysis of a larger sample of data, along with the addition of further daily-life activities that can be inferred from IMU. Moreover, we intend to conduct a longitudinal study to determine whether data acquired from the glasses could be used to track changes in the disease symptoms over time, which will be beneficial for inferring early disease progression or deterioration.

## Data Availability

All data produced in the present study are available upon reasonable request to the authors

https://emteq.net/

